# A Human Factors Approach to Childhood Pneumonia A qualitative study in Malawian primary care

**DOI:** 10.1101/2024.01.03.23300675

**Authors:** Balwani Mbakaya, Master R.O. Chisale, Tsung-Shu Joseph Wu, Mark T Ledwidge, Billy Nyambalo, Chris Watson, Joe Gallagher

## Abstract

**Introduction:** Pneumonia is one of the major causes of childhood mortality worldwide. Most of these deaths are readily preventable or treatable with proven cost effective interventions. The aim of this study is to investigate the assessment and management of childhood pneumonia in primary care in Malawi.

**Methods:** Semi-structured interviews were used to elicit accounts of the assessment, treatment and referral process as experienced by staff and caregivers in 10 health facilities in Northern Malawi. Staff members dealing with assessment of children under-five years of age (Doctors, Clinical Officers, Medical Assistants, Nurses, Nurse Midwife Technicians, Student Nurses and Health Surveillance Assistants) were included. It also involved policy makers from the area and caregivers of children presenting with pneumonia. Data was analysed using thematic analysis following the SEIPS 101 tools (task sequence, use of technology and tools, and work system barriers and facilitators).

**Results:** 43 interviews were undertaken. A process map of the flow of patients with pneumonia was created, showing the tasks undertaken and the interactions between staff and patients. In their interviews caregivers identified a number of barriers to appropriate care including insufficient education for caregivers on the management of sick children and distance to healthcare availability. Staff identified several organisational elements that served as barriers to the implementation of care. They included workload, lack of resources such as medications and batteries, uncertainty regarding markers of severity and need to give all antibiotics. The health passport system and effective teamwork and community were highlighted as being important facilitators.

**Conclusion:** This study provides information on the challenges and issues involved in managing childhood pneumonia in primary care in Malawi. These barriers included a lack of resources, staff and caregiver education and heavy workload. The ability to identify severe illness accurately and identify those who do not require antibiotics were highlighted as issues for health workers.

## Introduction

Despite recent advances in reducing childhood mortality, pneumonia remains a major cause of morbidity and mortality among children aged under five years of age(1) In primary care the focus is on providing antibiotics, identifying those children at risk of severe illness, and ensuring they are referred to hospital. The WHO has identified severity criteria to guide referral to hospital and these criteria have been implemented in initiatives such as Integrated Management of Childhood Illness (IMCI)(2). However recent studies have highlighted concerns regarding changing markers of severity which may mean children with severe illness may not be appropriately identified(3). Also WHO markers of severity, which are used to guide referral for hospitalization in primary care, are currently based on clinical symptoms alone but the wider availability of technology allows the use of markers such as pulse oximetry to be more widely integrated to help determine severity. A greater understanding and characterisation of these changes is required to continue advances in reducing childhood mortality from pneumonia. Most studies have focused on the hospitalized population, and yet in developing countries it is in the community where these patients are most likely to present(4).

The aim of this study were to establish the views of healthcare workers, policy makers and caregivers regarding the current process in the assessment and management of childhood pneumonia in primary care focusing on assessment of severity and decision to prescribe antimicrobials.

## Methods

This research was conducted at 10 health care facilities located in Northern Malawi and with policy makers in the Ministry of Health in Malawi in 2022. Semi-structured interviews were conducted to elicit in-depth accounts of the assessment process as experienced by staff and caregivers. This format was selected to ensure that there was enough structure to the interviews to allow them to be comparable but also allow enough flexibility to incorporate different staff levels without generating bias. Open questions were used to allow in-depth insight into the perspectives of participants. The SEIPS 101 framework was used to analyse them in order to highlight the system design in place and also how this design influences other connected processes within the clinic and referral to first level hospitals. This data, along with informal observations from the clinics, was used to construct a process map, detailing the flow of patients. Following this the tools in SEIPS 101 were used to identify gaps and possible solutions.

## Results

### Process mapping

Through interviews and informal observations around the facility a process map was developed. Figure 1. It shows the flow of patients through identifying illness at home, to community supports, attending the health surveillance assistant community health worker) and if necessary attending the hospital. It also shows involvement of informal health supports such as traditional healers, village elders and drug stores (informal pharmacies). Where applicable it shows tools used at each level. Fig. 1

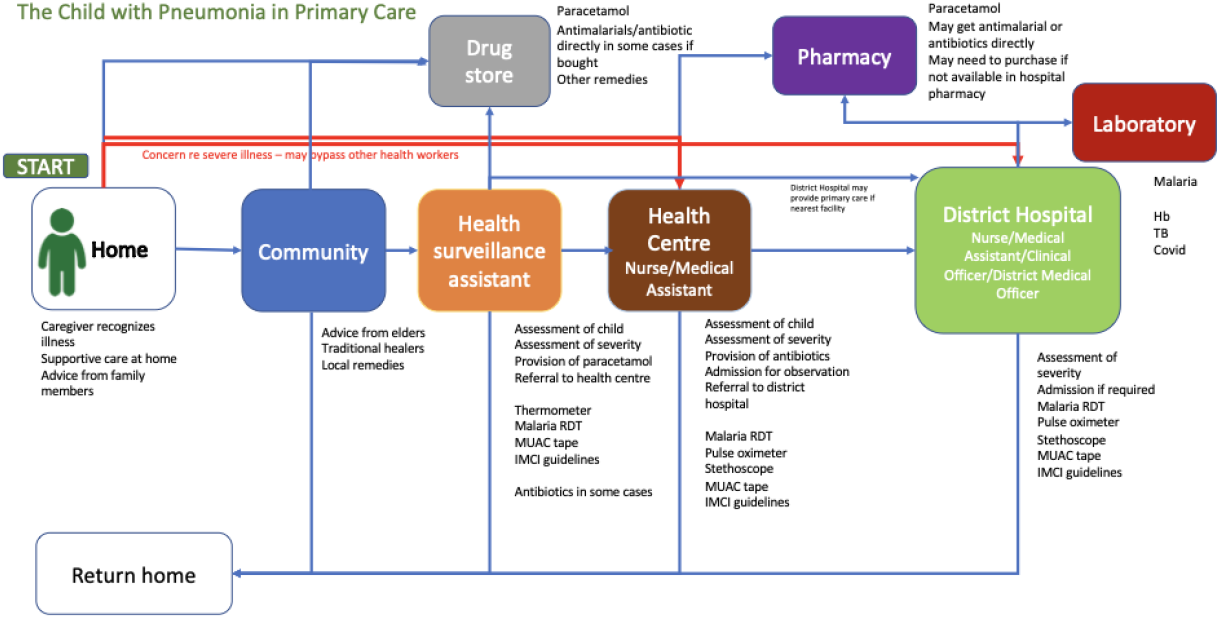

### Themes and sub-themes

The SEIPS 101 model was then used to explore the interviews. A PETT scan (People, Environment, Technology and Tasks) analysis was undertaken with barriers and facilitators highlighted. We also highlighted the tasks involved, tools used and outcome identified as important (supplemental file) The themes and subthemes are identified below. Following this local artists created a sculpture based on the experiences portrayed entitled “The Fight for Life” which demonstrates these findings in a visual manner (supplemental file)

#### Theme 1: The fight for life

During the interview process, guardians to children with pneumonia expressed several concerns regarding the struggle that they go through with their children to get help/treated once they get sick. The challenges identified were the decision to take the child to a health facility, choice of a health care service, and transport to a health facility. The major subthemes are outlined below

#### Sub-theme 1: Decision to take the child for health care

Mothers and guardians go through several steps to reach a decision to go with a child to a health facility or a traditional healer seeking advice from others.

*When the child is sick, I have to consult the husband or mother-in Law to decide when to go and where to seek health care*.

*I only consult when my husband is around. But most of the time I just decide to go alone because my husband is always away to do piece work and I do not have a phone to call him*

#### Sub-theme 2: Choice of health care service

When it come to health care seeking behaviour, specifically to decide whether to go to health facility or traditional healers, a number of guardians highlighted that they would go to traditional healer first before attending a health worker sometimes dues to belief but also due to cost.

*Most of the timewhen the child has breathing problems we frequently believe that wicked forces have been at work through witchcraft which is initiated by a family member or neighbour. Hence some traditional healers (sing’anga) can be able to unveil the culprit (kuombeza) and prescribe the appropriate medicine to counteract the threat to life*.

Another guardian had this to say;

*It is relatively cheaper at the traditional healer than at the health facility, and also hospital is far. Some traditional healers live with us in the same village*.

#### Sub-theme 3: Transport to go to the facilities

Many participants expressed concern about the challenges they face with transport to take their children to the hospital/health facility when they are sick. Some parents also expressed worry that they end up delaying to get treatment early because of struggles with transportation. Below are some of the quotes from the guardians;

*Sometimes I just walk on foot with the child at my back, but I take 4 hours to reach the nearest health centre*.

Another mother to a pneumonia child said;

*I hire a bicycle to take me to the hospital with my child*.

While the other mother said;

*I normally have to sell a chicken so that I use money for transport and also for treatment in case there are no drugs at the facility so I buy from a store*.

While the above theme and quotes reflect the stories from guardians, health workers during the interviews also expressed concerns and wished to have better support system through availability of tools to support their decisions; two main themes were developed. These are; (1) Indecisiveness, and (2) wish to better tools. Below are the themes and their quotes to support the theme;

#### Theme 2: Indecisiveness

Health workers who were interviewed, expressed concern over discomfort in making precise decision regarding severity of pneumonia and also prescription of the drugs; For example, one of the health workers said;

*At times I decide to admit the child not based on the flow charts on the wall because the child may not exactly fit into that, but looks sick*.

While the other health worker had the following to say;

*I do prescribe antibiotics just to cover the child even if it is not severe. After all that is what the guidelines tell us to do*

The other health worker said; *I hold the antibiotics and not give the child sometimes because of the MRDT (Malaria Rapid Diagnostic Test) results if positive*

Another health worker noted;

*I give antimalarial even if the child is negative because other types of plasmodium cannot be picked by the MRDT*.

#### Theme 3: Wish for better tools

When health workers were asked during the interview what they would want to assist them with precision of severity assessment and prescription of antibiotics, several different responses came out as follows. For example, most participants said would want to have the following to help with severity classification;

*Pneumonia mobile app*

*App integrated with clinical record/research App to collect anonymised data*

When they were asked the type of features to be included in the mobile app for Pneumonia assessment, the health workers mentioned the following;

*App should include signs and symptoms*

*App should contain standard guidelines with both internal and external controls*

*App should be accurate, friendly, simple, free, accessible, not dependent on internet*

*App should differentiate bacterial, viral and fungal pneumonia*

*App should have high sensitivity and specificity*

*App should not take long to give results*

The further emphasized that a tool to help differentiate viral from bacterial pneumonia in order to minimise use of antibiotics and prevent resistance would be of beneft and they felt would be widely used. It was proposed that the tool should be similar to how the malaria rapid diagnostic test in Malawi works which is a lateral flow test which uses a finger prick blood sample.

## Discussion

This needs assessment highlighted a number of issues with the assessment and management of childhood pneumonia and opportunities for intervention.

It highlighted the difficulties for parents and guardians in making a decision to seek care and who to seek care from as well as the financial implications of seeking care. Delays in seeking care are associated with worse outcomes and have been reported previously also(5). The prominent use of traditional healers in this study is different to previous studies. Previous studies have shown that of children who died in Malawi most caregivers brought their child to healthcare, and many sought care multiple times from different healthcare providers, suggesting issues in identifying children with severe pneumonia also(14). Quality of health services has been associated with better healthcare utilisation(6). Therefore a focus on improving quality of services and providing supports for healthworkers in terms of tools and resources is important. Although the majority of pneumonia episodes are non-severe and are managed in at primary healthcare facilities and therefore studies in primary care are vital(7). Although it may be presumed that if the illness worsens, the child may progress through the hierarchy of the healthcare system from primary to secondary or tertiary care other studies have shown that significant numbers of children die without ever attending hospital with one study showing that approximately two-thirds of children who die do so in the community (8). The WHO criteria for severe disease are used to guide referral to hospital from primary care. A study by our group in primary care in Malawi showed that WHO severity criteria were present in a minority (30.4%)(11) of children hospitalised, consistent with other recent work showing that 39% of fatal cases of pneumonia in children in hospital were defined as having non-severe pneumonia requiring only home treatment by the 2013 revision. The fact that the WHO criteria would have discharged these children with oral antibiotics highlights the need for new markers of severity, as these guidelines are used to guide primary care workers in their decision-making regarding escalation of care. Healthworkers in this study highlighted differences between their opinions of illness severity and what guidelines gave as severity markers. Developing dynamic systems of identifying changing patterns of risk will be important. Systems using analysis of routinely collected health data and artificial intelligence approaches may be an approach to this.

There is also growing concern regarding antimicrobial resistance driven by overuse of antibiotics and intercontinental spread of this resistance. The increased use of universal immunisation, including haemophilus influenzae type B (Hib) and pneumococcal vaccines has altered the proportion with viral illness(7,8). Current guidelines recommend that all children with WHO defined pneumonia receive antibiotics. However, a recent study has also shown that, in Malawi, the number of children with non-severe, fast-breathing pneumonia that needed amoxicillin treatment, for one child to benefit was 33(9). In order to reduce use of antibiotics point of care tests that are accurate in primary care and easily available in primary care are important. Similar approaches in malaria have yielded significant reductions in antimalarial prescription(10). The use of a test such as the malaria rapid diagnostic test was highlighted as a solution by health workers in this study.

This study had a number of limitations. Parents and guardians were recruited from health facilities and so were likely there with a sick child. This may have influenced opinions from the parents based on their recent experiences. A diverse cadre of staff were interviewed to provide different perspectives from the community to health policy. It may be beneficial to explore each role more thoroughly in future studies. The observations were also carried out within the healthcare facility by the researchers to assist in developing the SEIPS assessment. Although informal, the presence of researchers may have influenced how staff acted.

Overall the use of a diverse range of participants has allowed the researchers to identify the process, people, tools, tasks and outcomes for a child with pneumonia in the community. It highlights the areas where improvements are possible.

## Data Availability

All data produced in the present study are available upon reasonable request to the authors

## Acknowledgements

This study was funded by Science Foundation Ireland (SFI) and the Department of Foreign Affairs (DFA) under the SDG Challenge Grant Number SFI/21/FIP/SDG/9948

