## Supplemental file for "A Human Factors Approach to Childhood Pneumonia A qualitative study in Malawian primary care"

**Supplemental File:** **1**
**S1 Characteristics of sites** **1**
**S2 Participant characteristics** **1**
**S3 People, Environment, Tools, Tasks (PETT) scan** **1**
**S4 Tasks Matrix** **3**
**S5 Tools Matrix** **4**
**S6 Outcomes Matrix** **5**
**S7: Sculpture** **6**

#### S1 Characteristics of sites

| Site Name | Type |
|---|---|
| Bwengu | Health Centre |
| Bolero | Health Centre |
| Jenda | Health Centre |
| Mapale | Health Centre |
| Mpamba Road | Health Centre |
| Chinteche | Health Centre |
| Rumphi | District Hospital |
| Mzimba South | District Hospital |
| Nkhata Bay | District Hospital |
| Mzuzu | Central Hospital |

#### S2 Participant characteristics

| Type | Number |
|---|---|
| Parent/Guardian | 4 |
| Nurse Midwife Technician/Nurse/Midwife | 18 |
| Medical assistant/Ciinical Officer | 12 |

| | |
|---|---|
| Doctor | 2 |
| Policy maker | 7 |
| Lab Technologist | 13 |
| Health Surveillance Assistant | 9 |

#### S3 People, Environment, Tools, Tasks (PETT) scan

| **Factor** | **Group** | **Barriers** | **Facilitators** |
|---|---|---|---|
| People | · Child<br>· Parent/Guardian<br>· Local healer<br>· HSA<br>· Nurse<br>· Medical assistant<br>· Clinical officer<br>· Other family members e.g. grandparents for advice<br>· Others | Inadequate civic education on what to do when a child has a cough | Availability of some health services |

| | | | |
|---|---|---|---|
| Environment | · Home<br>· Village<br>· Health Centre<br>· District Hospital<br>· Tertiary Hospital<br>· District Health Office<br>· Ministry of Health<br>· CHAM<br>· Others | Distance to health facility,<br>Delay in seeking health services for a pneumonia child<br>Workload<br>Availability of resources<br>Time management<br>Transportation<br>Space<br>Staff motivations<br>Confidence<br>Family expections<br>Cost<br>Patient education | Effective communication and collaboration among different stakeholders<br>Training<br>Availability of health passport<br>Teamwork<br>Prioritisation |
| Tools | · Guidelines<br>· Education materials<br>· Stethoscope<br>· Pulse oximeter<br>· Malaria RDT<br>· Antibiotics<br>· Others . . . | Not all skilled in effectively using stethoscope<br><br>Pulse oximeter not always functional due to<br>Batteries<br><br>Guidelines not available | Readily availability of WHO (IMCI) guidelines |

| | | | |
|---|---|---|---|
| Tasks | Identify child is sick<br>Treat at home first with local remedies?<br>Get advice locally from who<br>Attend health professional<br>History from parents<br>Exam using thermometer, pulse oximeter, stethoscope, malaria RDT<br>Decide on cause of illness<br>Decide if needs treatment<br>Decide what treatment<br>Decide if needs higher level of care<br>Explain to parents<br>Obtain treatment and advise on its use<br>Advise when to come back | Inadequate resources to use for diagnosis of pneumonia and treatment/management approach<br><br>Personal judgement and experience | Accessibility to health facility and services |

**Table 1 PETT scan**

### S4 Tasks Matrix

**Tasks Matrix**

| | **Who does it** | **Goal of task** | **Frequency** | **How performed** | **When performed** |
|---|---|---|---|---|---|
| Identify child is sick | Parent | To see if unwell and if needs treatment | Always | Listens to child<br>Feels if hot<br>Sees if not feeding | At home when child coughing, short of breath, not feeding |

| | | | | | |
|---|---|---|---|---|---|
| Making a diagnosis of pneumonia | Nurses, clinicians, HSAs | To confirm if a child has pneumonia | Every encounter with a child presenting with cough | Use WHO guidelines, IMCI, stethoscope | When a child has a cough and visit the health facility |
| Treating a child with pneumonia | Nurses, clinicians, HSAs | To cure a child with pneumonia | Every pneumonia diagnosis | Use WHO guidelines, IMCI, stethoscope | When a child has a cough and visit the health facility |

### S5 Tools Matrix

**Tools matrix**

| | Users | Purpose of use | Frequency of use | Ease of access | Usability |
|---|---|---|---|---|---|
| Stethoscope | Nurse, medical assistant, clinical officer | To determine if extra lung sounds | When child is with these health professional | Found in health centre, hospital only | Needs training, can be hard to hear if child crying |
| IMCI guidelines | Nurses, clinicians, HSA | To guide in reaching pneumonia diagnosis | When child is with these health professional | Found in health centre, hospital only | Needs training |
| Pulse oximeter | Nurses, clinicians, HSA | To determine the breathing rate (fast breathing) | When child is with these health professional | Found in health centre, hospital only | Needs training |
| Antibiotics | Nurses, clinicians, HSA | To treat a child with pneumonia | When a child is diagnosed with pneumonia | Found in health centre, hospital, pharmacy | Needs training on prescription |

### S6 Outcomes Matrix

**Outcomes matrix**

| | | | |
|---|---|---|---|
| | **Child/family** | **Healthcare professionals** | **Organisation** |
| *Desirable* | Receive antibiotics appropriately<br>Refer to higher level of care appropriately<br>Receive care near home if possible | Accurate diagnosis<br>Timely diagnosis<br>Appropriate workload<br>Appropriate tools | Appropriate level of care<br>Cost of process |
| *Undesirable* | In appropriate treatment | Inaccurate diagnosis | Overwhelmed system |
| | Delayed referral to the next level of care | Inappropriate referral system | |
| | Long distance to receive care | Overwhelming responsibilities/ tasks | |

S7: Sculpture

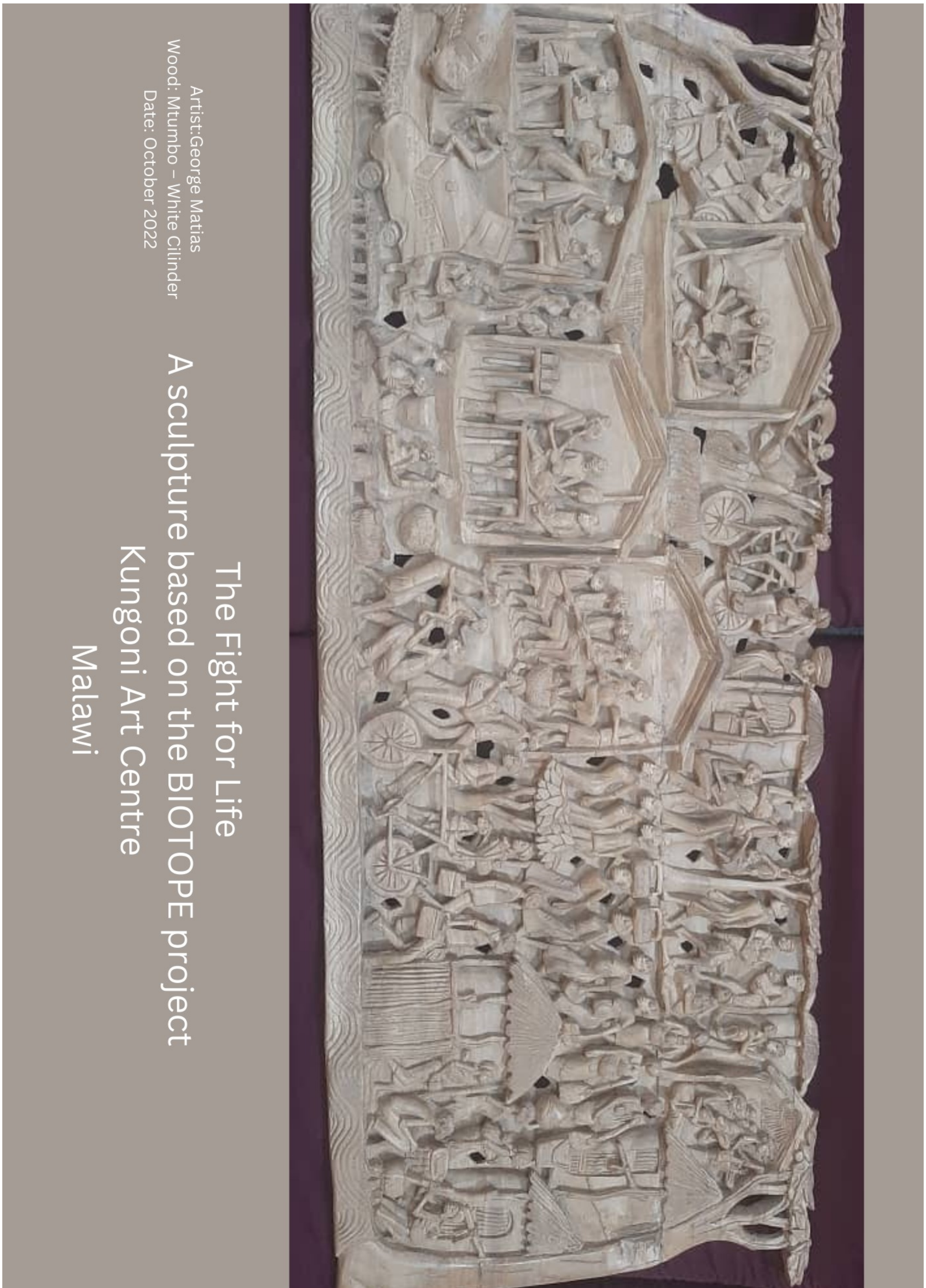

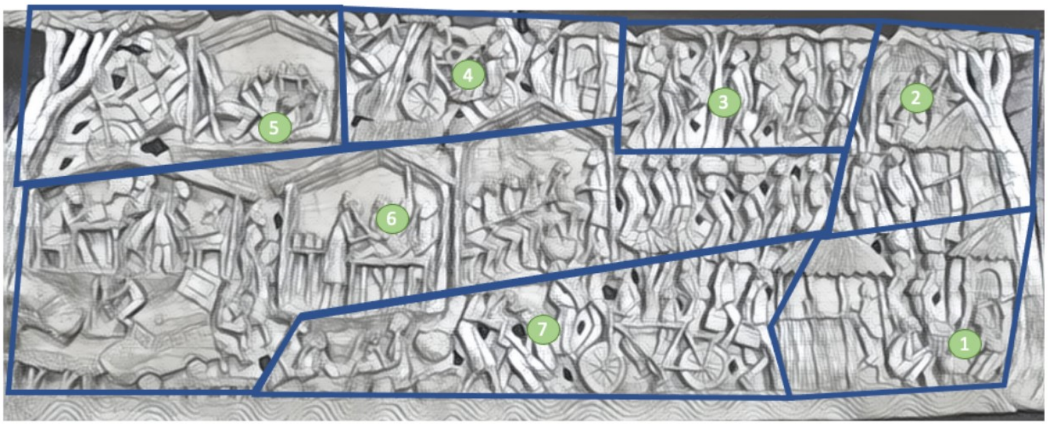


**1. Family Discussions**
The family has started to gather as the child is showing signs that there is something wrong health wise. The mother will be the first to notice any such changes to the health and body temperature of the child. She will then begin to ask advice from her mother, sisters or grandmother. This context is specific to the Chewa where the woman is responsible for the primary care of the child. In a patrilineal setting such as the Tumbuka it would be the husbands sisters and mother who would determine what medical assistance should be provided for the child. Either way the women will be central to any such decisions.
A large part of the discussion will depend on how much money is available at that time. There is often no savings from which they can access some money. It is for this reason that the first step in seeking medical help is often protracted and adds to the emerging sickness. Some may initially just buy some tablets i.e. paracetamol at the local shop and hope for the best.
The decision as to what medial practitioner they will utilise also depends on the educational background of the parents. Some may be much more familiar with local medicine than attending the larger hospitals. Distance also plays a large part in where to seek help. Since the majority of people have no personal transport it means extra costs will be incurred which may not be possible for the family. Therefore, the first port of call will be the traditional healer (Sing'anga)

**2. The Traditional Healer – Mankhwala ya Chikuda**
On the panel we see the women walking to the place of the sing'anga. This may take some time as the place may not be in their village. However, walking means that some money is saved and can be used for the services they will avail of. Some practitioners will also accept chickens or maize in the place of money. This will suit many people as their access to available cash is often limited but they can easily take some maize from the store or catch a chicken. Sadly, this is often the case with many emergencies which come to the homestead and it seriously effects the food security of the family.
The traditional healers are varied in their capacity and expertise. Some may focus exclusively on the use of leaves, roots and seeds and use the knowledge they have received from their elders to mix and administer. In the panel we see the "doctor" pounding his mixture in a mortar. However, the arrival of sickness into a home is surrounded by many different understandings. For some it seems incomprehensible that the health of a family member can be taken so quickly by natural causes. There is frequently a belief that malevolent forces have been at work through witchcraft which is initiated by a family member or neighbour. Some sing'anga claim to be able to unveil the culprit (kuombeza) and prescribe the appropriate medicine to counteract the threat to life. It may involve washing the child in medicated water and the wearing of medicinal strings and amulets.  This may be protracted and merely add the sickness of the child. If there is no change then the next option is to enter into the formal medical process.

**3. First Contact - Za Umoyo**

In all villages there are trained Health Surveillance Assistants (HSA) who are considered the first contact for someone to enter the formal medical system. The officials also supervise the general health of the village taking notes of water sources and sanitary standards. They live in the village themselves and coordinate with the health centre. We can see one person writing the medical details in the Health Passport, which will be a reference point for the impending visit to the health centre or hospital. These assistants may also distribute some medicine to children under the age of five for sickness such as malaria or diarrhoea.

In this panel on top we see the children being weighed and assessed for nutritional problems. This may take place in a central place in the village under a large tree or some villages may have built a small building to help in this process. When such gatherings are planned they are announced through the chiefs and also through the churches on their chosen day of prayer. While traditionally it was only women who attended "scalu" it is now encouraged that the father of the child is also present.

**4. Getting from A to B – Modes of transport**

Transport is a serious consideration in any medical journey. Some may have their own means but for many they will rely on a relative or hire a local means. Here the father of the child will have to play a role in organising the arrangements. At the top of the panel we see the mother being brought by bicycle to the health centre. As mentioned the cyclists may be the father, a family relative or someone who is hired (dumper). Previously, there was a bike given to the village which could be used by people freely to access medical help but this has now stopped.

As they travel on their journey they pass someone busy in their garden hoping to make some gains from the sale of his produce. He may also be thinking of cutting the tree to planks of wood or burning for charcoal. Others rely on casual work (ganyu) or the sale of small animals. It is these efforts which will provide access to money even at a very minimum level. People do not usually save money for a medical emergency but focus on very short term needs. This means an issue of transport and the type being used can seriously influence how a sickness is going to be dealt with.

**5. Help Nearby- The Local Health Centre**

Towards the top right hand corner, at the local health centre, the child will be received by a clinician or medical assistant. There is usually no more than one available to help people. The Health Passport will be consulted to understand the medical history of the child. This written record proves invaluable in assessing the progress of a sickness and the health status of the child. The services provided at the health centre are minimal although they have the capacity to admit and monitor the situation. They may give some medicine to reduce such symptoms as fever. If the situation deteriorates then they will be referred to the District hospital. It depends how critical the sickness become will determine the mode of transport to be used. An emergency will require the call for an ambulance which is provided freely by the health system. Otherwise the parents will be forced to seek another means of transport. Since the distance can be long and the sickness is progressing they may hire a motor bike (kabaza). We should note that the mother of a child would not deal with this situation on her own but would be accompanied by her relatives. They would also need to travel adding to the expenses of the situation.

**6. More Choice - District Hospital**

The arrival at the District Hospital is depicted in three scenes on the left, each with a roofed building. The number of trained personnel increases at this level with one of two doctors and more clinicians. The first station shows people arriving by ambulance. Someone is busy washing his hands remembering the need for hygiene and preventing the spread of more sickness. If the child is seriously sick they would be received by a nurse as shown in the panel. The use of masks is still considered a priority to protect all those used the health services. The child will be brought to the first station in order to once more be assessed through consultation of the medical passport and checking the vital signs. The health official can see form the written record the journey the child has made form the initial contact at the village level. If need be the child will be brought to the second station of paediatrics ward where a more in-depth assessment can be made by a doctor. In this scene we see the examination in progress and a drip has already been attached in order to restore some strength. It is here that the full diagnosis will be determined and the appropriate care would be agree. If it is pneumonia (zibayo) the treatment will follow. In the centre there is a line of women making their way to the hospital so they can also get the help needed especially for pre-natal care.

**7. Home at Last – Follow up**

At the bottom of the panel we see a much happier scene where the mother and father enjoy the presence of their healthy child. We see relatives coming the visit and have an update on how everything went at the hospital. The follow up care of the child is very important and we see the local health assistants returning after a heath visit to the family concerned. Such visits are irregular due to the poor number of trained personnel. However, it is often the advice given by the family members which will determine how much care will be taken to ensure the child has a healthy life.
